# Burden and prevalence of prognostic factors for severe covid-19 disease in sweden

**DOI:** 10.1101/2020.04.08.20057919

**Authors:** Katalin Gémes, Mats Talbäck, Karin Modig, Anders Ahlbom, Anita Berglund, Maria Feychting, Anthony A. Matthews

## Abstract

**Objectives:** Describe the burden and prevalence of prognostic factors of severe COVID-19 disease at national and county level in Sweden.

**Design:** Cross sectional study

**Setting:** Sweden

**Participants:** 9,624,428 individuals living in Sweden on 31st December 2014 and alive on 1st January 2016

**Main outcome measures:** Burden and prevalence of prognostic factors for severe COVID-19 based on the guidelines from the World Health Organization and European Centre for Disease Prevention and Control, which are age 70 years and older, cardiovascular disease, cancer, chronic obstructive pulmonary disease, severe asthma, and diabetes. Prognostic factors were identified based on records for three years before 1^st^ January 2016 from the Swedish National Inpatient and Outpatient Specialist Care Register, Prescribed Drug Register, and Cancer Register.

**Results:** 22.1% of the study population had at least one prognostic factor for severe COVID-19 (2,131,319 individuals), and 1.6% had at least three factors (154,746 individuals). The prevalence of underlying medical conditions in the whole study population ranged from 0.8% with chronic obstructive pulmonary disease (78,516 individuals) to 7.4% with cardiovascular disease (708,090 individuals), and the county specific prevalence of at least one prognostic factor ranged from 19.2% in Stockholm (416,988 individuals) to 25.9% in Kalmar (60,005 individuals).

**Conclusions:** The prevalence of prognostic factors for severe COVID-19 disease will aid authorities in optimally planning healthcare resources during the ongoing pandemic. Results can also be applied to underlying assumptions of disease burden in modelling efforts to support COVID-19 planning. This information is crucial when deciding appropriate strategies to mitigate the pandemic and reduce both the direct mortality burden from the disease itself, and the indirect mortality burden from potentially overwhelmed health systems.

## INTRODUCTION

As of 8^th^ April 2020, the number of confirmed COVID-19 infections surpassed 1.3 million worldwide, and the number of infections leading to death reached 74,000.[1] Guidelines from the World Health Organization and the European Centre for Disease Prevention and Control suggest that individuals aged 70 years and older or with an underlying medical condition such as cardiovascular disease, high blood pressure, cancer, chronic obstructive compulsory disease (COPD), asthma, and diabetes, are considered to be at high risk of developing severe disease, and hence may require critical care.[1, 2] These recommendations are mainly based on studies from the Wuhan region of China and Italy, and generally show that once infected, individuals with at least one of these prognostic factors are more likely to go on to generate severe disease and a resulting higher risk of mortality.[3-8] Governments around the world have, therefore, recommended that individuals with at least one of these factors self-isolate for prolonged periods of time to not only reduce the risk contracting severe COVID-19, but also prevent any sudden increase in demand for critical care in hospitals, which could overwhelm health systems. If the pandemic developed to affect a large proportion of the population, then critical care capacity could become saturated. However, the prevalence of these prognostic factors for severe disease are to a large extent unknown in many countries. Knowledge of the distribution of individuals considered to be at high risk of severe COVID-19 disease, coupled with the capacity of the health care system, would allow clear strategic planning.

Several models have been produced to support COVID-19 planning in countries across the world.[9-12] Many of these models are based on the assumption that disease severity increases with age, but they do not account for an increased risk of severe disease in individuals with underlying medical conditions. This is usually because age stratified burden of disease at a local level is rarely available. Even when this information is available, data from which it originates can be obtained from a sample of the population rather than from the whole population. If the sample is not representative of the population at large, results may be biased. In order to build clear robust models that will provide trustworthy estimates of the extent to which the infection will impact populations, we need reliable estimates on the underlying prevalence of medical conditions suggesting high risk of severe disease.

The unified Swedish healthcare and register system provides a unique opportunity to calculate the burden and prevalence of prognostic factors for severe COVID-19 disease. This knowledge will help both healthcare capacity planning and provide further data that can be applied to underlying assumptions for models that support planning worldwide. We therefore aimed to use Swedish register data to describe the prevalence of prognostic factors of severe COVID-19 disease at national and county level in Sweden.

## METHODS

### Data sources

We used data from the Swedish national health care and population registers linked at an individual level using the unique personal identification number of all residents in Sweden.[13] Disease burden was based on diagnoses and date of hospitalization or visits from the National Inpatient Register and Outpatient Specialist Care Register, and sociodemographic characteristics such as age, sex, county of residence were obtained from the Total Population Register.[14, 15] We also used the Cancer Register to identify malignant tumors, and the Swedish Prescribed Drug Register to identify prescriptions dispensed by individuals and further our understating of disease burden.[16, 17] These data were originally aggregated as part of a study on comorbidities in cancer risk and survival.

### Study population

We identified all people living in Sweden on 31^st^ December 2014 and alive on 1^st^ January 2016.

### Identification of prognostic factors for severe COVID-19 disease

We based our decision on the prognostic factors for severe COVID-19 on the guidelines from the World Health Organization and European Centre for Disease Prevention and Control,[1, 2] which were age 70 years and older, cardiovascular disease, cancer, COPD, severe asthma, and diabetes. Age was calculated at 31^st^ December 2015. An individual was then identified as having an underlying medical condition if they had a diagnosis in either the Inpatient or Outpatient Register (as primary or secondary diagnosis) or the Cancer Register within three years prior to 1^st^ January 2016. If data were available, we also identified related dispensations of prescriptions from the Prescribed Drug Register within the same time period. We used International Statistical Classification of Diseases and Related Health Problems version 10 codes (ICD-10) to identify a diagnosis and Active Therapeutic Chemical codes (ATC) to identify the dispensation of a prescription. The ICD-10 and ATC codes for underlying medical conditions were: cardiovascular disease (I20-I99), cancer (C00-C75), COPD (J41-J44), severe asthma (J45), and diabetes (E10, E11, E13, E14, O24; ATC: A10).

### Analysis

We initially calculated the burden (raw number) and prevalence (proportion) of all individuals living in Sweden in relation to their sex, age (1-9, 10-19, 20-29, 30-39, 40-49, 50-59, 60-69, 70-79, 80+), county of residence (21 counties), if they had a predetermined prognostic factors for severe COVID-19, and if they had at least one, two, or three of these prognostic factors. We then calculated the burden and prevalence of the five underlying medical conditions across each age group, and the burden and prevalence of each of the six prognostic factors individually in each of the 21 counties across the whole of Sweden. We also calculated the age group stratified burden and prevalence of each of the five underlying medical conditions in each county. Finally, we calculated the burden and prevalence of individuals with at least one, two, and three prognostic factors for severe COVID-19 in each county.

### Sensitivity analyses

We repeated all above analyses using a look back period of one, five and ten years prior to 1^st^ January 2016 to define occurrence of disease in the registers, rather than three years.

### Patient Involvement

No patients were involved in setting the research question or the outcome choices, nor were they involved in developing plans for design or implementation of the study. No patients were asked to advise on interpretation or writing up of results.

## RESULTS

Table 1 shows the characteristics of the study population. The mean age was 41 years and around 50% of the 9.6 million individuals lived in three of the 21 counties (Stockholm, Västra Götaland and Skåne). Over 22% had at least one prognostic factor for severe COVID-19 (2,131,319 individuals), and 1.6% had at least three factors (154,746 individuals).

**Table 1.** Characteristics of the study population*

| Characteristics | N (%) |
| --- | --- |
| **Total** | 9,624,428 (100.0) |
| **Female** | 4,814,553 (50.0) |
| **Age (yrs)** |  |
| 1-9 | 1,041,469 (10.8) |
| 10-19 | 1,046,237 (10.9) |
| 20-29 | 1,306,883 (13.6) |
| 30-39 | 1,203,053 (12.5) |
| 40-49 | 1,302,755 (13.5) |
| 50-59 | 1,220,303 (12.7) |
| 60-69 | 1,146,021 (11.9) |
| 70-79 | 856,218 (8.9) |
| 80+ | 501,489 (5.2) |
| Mean age (yrs) (SD) | 41.4 (23.6) |
| **County** |  |
| Stockholm | 2,174,039 (22.6) |
| Uppsala | 346,325 (3.6) |
| Södermanland | 277,220 (2.9) |
| Östergötland | 436,474 (4.5) |
| Jönköping | 339,444 (3.5) |
| Kronoberg | 186,374 (1.9) |
| Kalmar | 231,956 (2.4) |
| Gotland | 56,639 (0.6) |
| Blekinge | 152,136 (1.6) |
| Skåne | 1,272,311 (13.2) |
| Halland | 308,576 (3.2) |
| Västra Götaland | 1,612,184 (16.8) |
| Värmland | 270,128 (2.8) |
| Örebro | 284,574 (3.0) |
| Västmanland | 258,458 (2.7) |
| Dalarna | 274,870 (2.9) |
| Gävleborg | 275,551 (2.9) |
| Västernorrland | 238,772 (2.5) |
| Jämtland | 124,686 (1.3) |
| Västerbotten | 258,371 (2.7) |
| Norrbottn | 245,340 (2.5) |
| **Prognostic factors for severe COVID-19** |  |
| Age 70+ years | 1,357,707 (14.1) |
| Cardiovascular disease | 708,090 (7.4) |
| Cancer | 129,155 (1.3) |
| COPD | 78,516 (0.8) |
| Severe asthma | 215,793 (2.2) |
| Diabetes | 471,015 (4.9) |
| **Number of prognostic factors for severe COVID-19** |  |
| At least one | 2,131,319 (22.1) |
| At least two | 649,935 (6.8) |
| At least three | 154,746 (1.6) |

\*On 1<sup>st</sup> January 2016

### Burden and prevalence of underlying medical conditions suggesting high risk severe COVID-19 by age group

Table 2 shows the burden and prevalence of each medical condition by age group in the study population. The prevalence of each condition generally increased as age increased; however, there was a higher prevalence of severe asthma in the youngest groups compared with all other age groups.

**Table 2.**
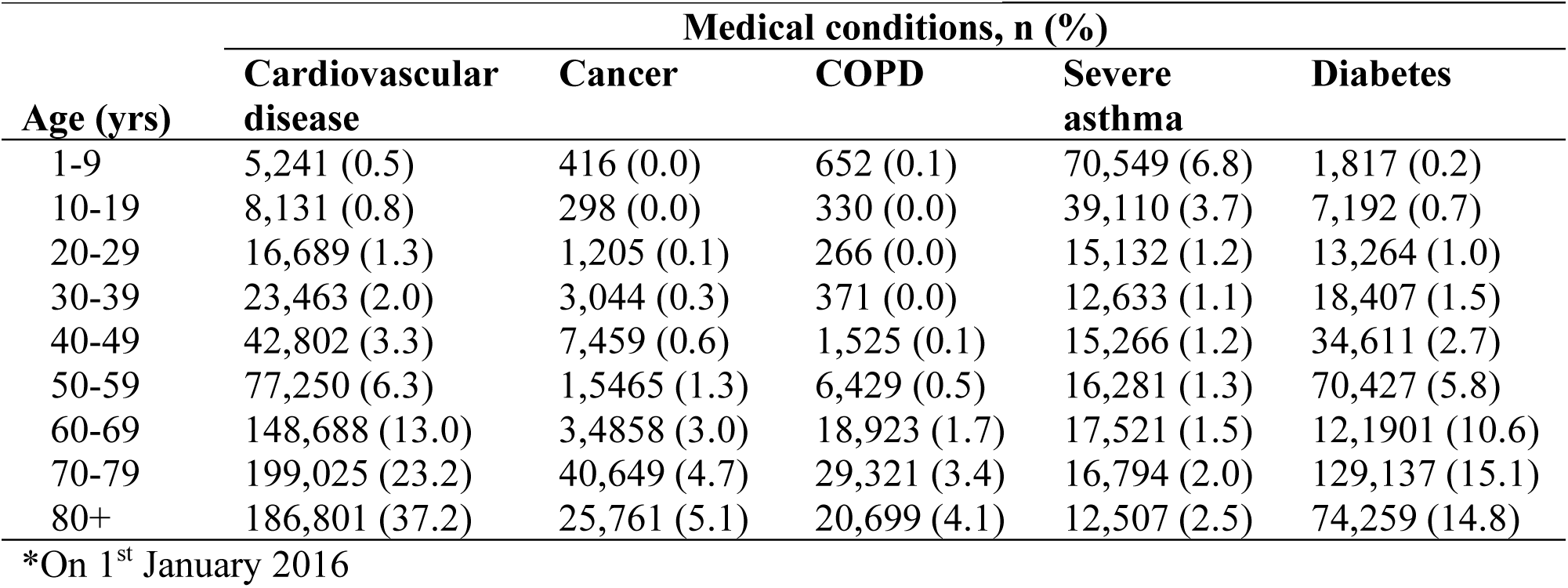
Burden and prevalence of underlying medical conditions suggesting high risk for severe COVID-19 by age group*

### Burden and prevalence of prognostic factors for severe COVID-19 by Swedish county

Table 3 shows the burden and prevalence of each of the six prognostic factors in each Swedish county, and Figure 1 visualizes the ratio of the county specific prevalence of each factor to the prevalence of that factor overall in the study population. The burden and prevalence of all prognostic factors were as follows (Table 3): the proportion of people aged 70 years and older ranged from 11.1% in Stockholm (242,208 individuals) to 17.6% in Kalmar (40,872 individuals); cardiovascular disease ranged from 6.4% in Uppsala and Stockholm (22,086 and 140,165 individuals) to 8.8% in Dalarna (24,205 individuals); cancer ranged from 1.1% in Norrbotten (2,790 individuals) to 1.6% in Halland (5,067 individuals); COPD ranged from 0.6% in Västerbotten (1,708 individuals) to 1.1% in Kalmar (2,448 individuals); severe asthma ranged from 1.8% in Västra Götaland and Örebro (29,353 and 5,243 individuals) to 2.6% in Norrbotten and Stockholm (6,407 and 56,251 individuals); and diabetes ranged from 4.0% in Stockholm (86,195) to 4.7% in Uppsala (16,311). The burden of each of the five underlying medical conditions stratified by age group in each county are also presented in Appendix 1.

**Table 3.** Burden and prevalence of prognostic factors for severe COVID-19 by each Swedish county*

| County | Prognostic factors, n (%) |  |  |  |  |  |
| --- | --- | --- | --- | --- | --- | --- |
|  | Age 70+ years | Cardiovascular disease | Cancer | COPD | Severe asthma | Diabetes |
| Stockholm | 242,208 (11.1) | 140,165 (6.4) | 27,234 (1.3) | 16,729 (0.8) | 56,251 (2.6) | 86,195 (4.0) |
| Uppsala | 44,294 (12.8) | 22,086 (6.4) | 4,328 (1.2) | 2,615 (0.8) | 8,193 (2.4) | 16,311 (4.7) |
| Södermanland | 43,914 (15.8) | 20,608 (7.4) | 3,470 (1.3) | 2,923 (1.1) | 5,861 (2.1) | 15,119 (5.5) |
| Östergötland | 63,485 (14.5) | 32,390 (7.4) | 6,228 (1.4) | 3,766 (0.9) | 9,807 (2.2) | 21,929 (5.0) |
| Jönköping | 50,850 (15) | 26,994 (8.0) | 4,738 (1.4) | 3,295 (1.0) | 8,121 (2.4) | 18,028 (5.3) |
| Kronoberg | 28,654 (15.4) | 13,483 (7.2) | 2,640 (1.4) | 1,704 (0.9) | 4,608 (2.5) | 9,272 (5.0) |
| Kalmar | 40,872 (17.6) | 20,270 (8.7) | 3,570 (1.5) | 2,448 (1.1) | 4,782 (2.1) | 13,347 (5.8) |
| Gotland | 9,743 (17.2) | 4,809 (8.5) | 892 (1.6) | 549 (1.0) | 1,219 (2.2) | 3,022 (5.3) |
| Blekinge | 26,092 (17.2) | 12,389 (8.1) | 2,358 (1.5) | 1,435 (0.9) | 3,257 (2.1) | 8,080 (5.3) |
| Skåne | 178,470 (14) | 98,919 (7.8) | 17,844 (1.4) | 11,791 (0.9) | 27,964 (2.2) | 64,259 (5.1) |
| Halland | 47,199 (15.3) | 23,619 (7.7) | 5,067 (1.6) | 2,276 (0.7) | 7,300 (2.4) | 13,653 (4.4) |
| Västra Götaland | 221,567 (13.7) | 111,920 (6.9) | 21,510 (1.3) | 11,545 (0.7) | 29,353 (1.8) | 75,352 (4.7) |
| Värmland | 45,646 (16.9) | 22,693 (8.4) | 3,959 (1.5) | 2,189 (0.8) | 6,048 (2.2) | 16,784 (6.2) |
| Örebro | 42,958 (15.1) | 20,344 (7.1) | 3,563 (1.3) | 2,039 (0.7) | 5,243 (1.8) | 14,831 (5.2) |
| Västmanland | 40,335 (15.6) | 21,114 (8.2) | 3,596 (1.4) | 2,244 (0.9) | 5,446 (2.1) | 14,240 (5.5) |
| Dalarna | 46,436 (16.9) | 24,205 (8.8) | 3,533 (1.3) | 2,133 (0.8) | 6,512 (2.4) | 16,283 (5.9) |
| Gävleborg | 45,805 (16.6) | 23,572 (8.6) | 3,861 (1.4) | 2,299 (0.8) | 6,353 (2.3) | 16,031 (5.8) |
| Västernorrland | 40,340 (16.9) | 19,492 (8.2) | 3,159 (1.3) | 1,694 (0.7) | 4,764 (2.0) | 14,545 (6.1) |
| Jämtland | 20,373 (16.3) | 8,892 (7.1) | 1,717 (1.4) | 1,019 (0.8) | 2,485 (2.0) | 7,154 (5.7) |
| Västerbotten | 38,120 (14.8) | 19,337 (7.5) | 3,098 (1.2) | 1,708 (0.6) | 5,819 (2.3) | 12,675 (4.9) |
| Norrbotten | 40,346 (16.4) | 20,789 (8.5) | 2,790 (1.1) | 2,115 (0.9) | 6,407 (2.6) | 13,905 (5.7) |

\* On 1<sup>st</sup> of January 2016

**Figure 1.**
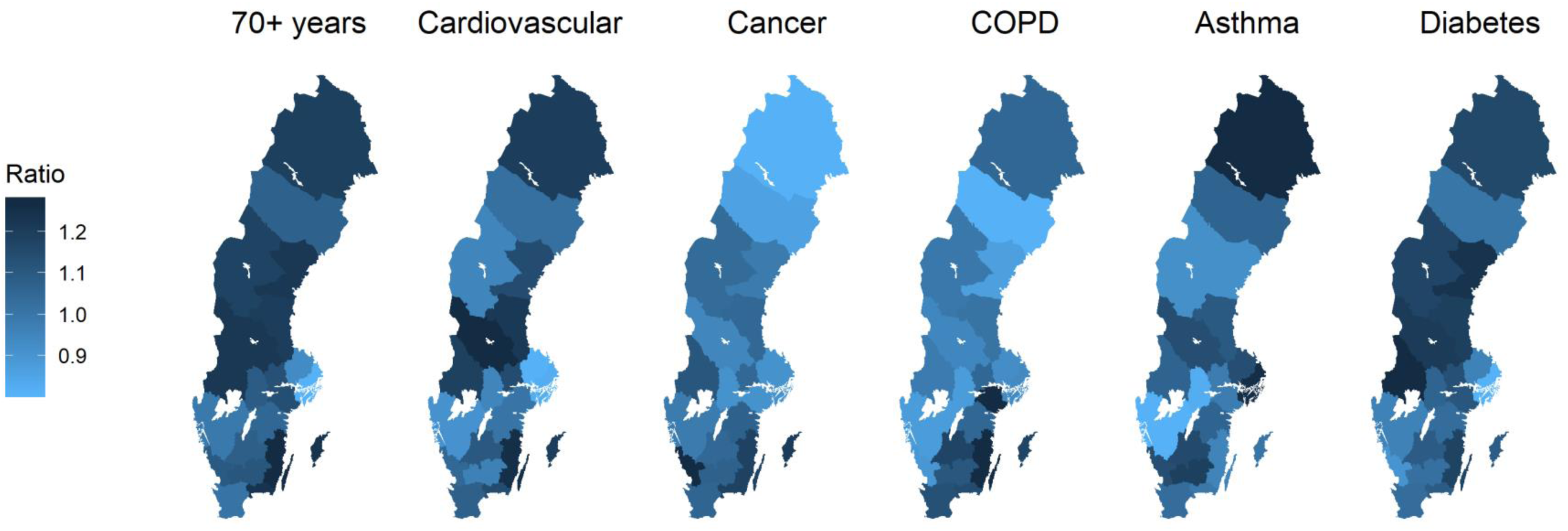
Maps showing the ratio of the county specific prevalence of each prognostic factor for severe COVID-19 compared with the overall prevalence of that factor in Sweden *Ratio corresponds to the country specific prevalence of each factor to the prevalence of that factor overall in the study population

### Burden and prevalence of at least one, two, and three prognostic factors for severe COVID-19 by Swedish county

Table 4 shows the burden and prevalence of individuals with at least one, two, and three of the six prognostic factors for severe COVID-19 in each county in Sweden, and Figure 2 visualizes the ratio of the county specific prevalence of people living with at least one, two, and three factors compared with the overall prevalence in the study population. The burden and prevalence of prognostic factors were: at least one prognostic factor ranged from 19.2% in Stockholm (416,988 individuals) to 25.9% in Kalmar (60,005 individuals); at least two prognostic factors ranged from 5.5% in Stockholm (119,057 individuals) to 8.5% in Kalmar (19,699 individuals; and at least three prognostic factors ranged from 1.3% in Stockholm (28,162 individuals) to 2.1% in Kalmar (4,839 individuals).

**Table 4.** Burden and prevalence of at least one, two, or three prognostic factors for severe COVID-19 in each Swedish county*

| County | Prognostic factors, n (%) |  |  |
| --- | --- | --- | --- |
|  | At least one | At least two | At least three |
| Stockholm | 416,988 (19.2) | 119,057 (5.5) | 28,162 (1.3) |
| Uppsala | 71,281 (20.6) | 20,674 (6) | 5,046 (1.5) |
| Södermanland | 66,330 (23.9) | 19,968 (7.2) | 4,786 (1.7) |
| Östergötland | 98,359 (22.5) | 30,431 (7.0) | 7,480 (1.7) |
| Jönköping | 78,639 (23.2) | 25,567 (7.5) | 6,634 (2.0) |
| Kronoberg | 43,911 (23.6) | 12,985 (7) | 3,005 (1.6) |
| Kalmar | 60,005 (25.9) | 19,699 (8.5) | 4,839 (2.1) |
| Gotland | 14,621 (25.8) | 4,468 (7.9) | 1,000 (1.8) |
| Blekinge | 38,357 (25.2) | 11,947 (7.9) | 2,832 (1.9) |
| Skåne | 285,175 (22.4) | 89,187 (7.0) | 21,485 (1.7) |
| Halland | 71,469 (23.2) | 21,780 (7.1) | 5,062 (1.6) |
| Västra Götaland | 341,102 (21.2) | 102,862 (6.4) | 23,704 (1.5) |
| Värmland | 68,976 (25.5) | 22,231 (8.2) | 5,350 (2.0) |
| Örebro | 65,005 (22.8) | 19,142 (6.7) | 4,288 (1.5) |
| Västmanland | 61,821 (23.9) | 19,586 (7.6) | 4,795 (1.9) |
| Dalarna | 70,688 (25.7) | 22,431 (8.2) | 5,238 (1.9) |
| Gävleborg | 69,341 (25.2) | 22,299 (8.1) | 5,448 (2.0) |
| Västernorrland | 60,339 (25.3) | 18,841 (7.9) | 4,266 (1.8) |
| Jämtland | 29,989 (24.1) | 9,175 (7.4) | 2,147 (1.7) |
| Västerbotten | 58,062 (22.5) | 17,906 (6.9) | 4,184 (1.6) |
| Norrbottn | 60,861 (24.8) | 19,699 (8.0) | 4,995 (2.0) |

\* On 1<sup>st</sup> of January 2016

**Figure 2.**
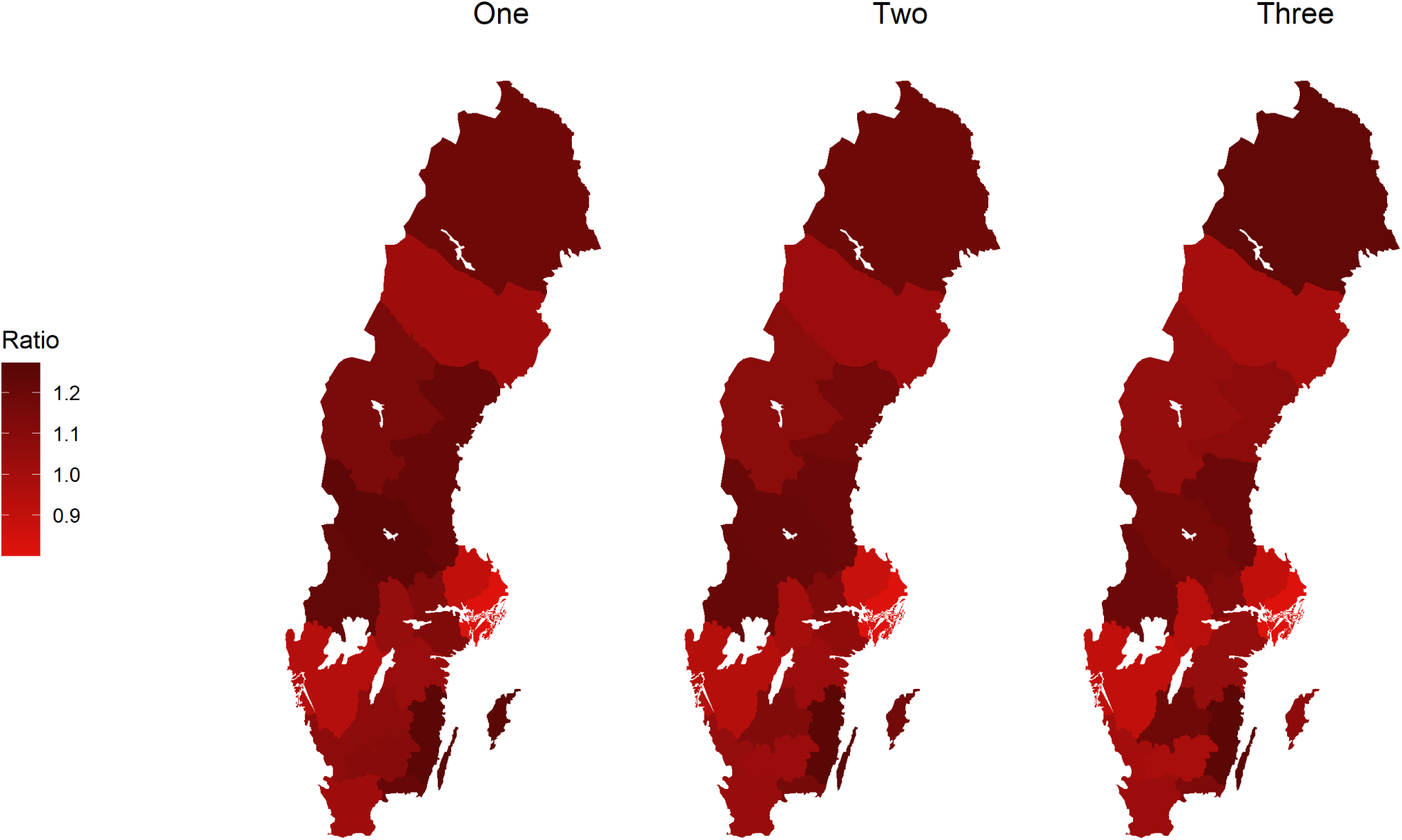
Maps showing the ratio of the county specific prevalence of people living with at least one, two, and three prognostic factors for severe COVID-19 compared with the overall prevalence in Sweden * Ratio corresponds to the country specific prevalence of people living with at least one, two, and three factors compared with the overall prevalence in the study population

### Sensitivity analyses

All analyses with a look back period of one, five, and ten years are shown in Appendices 2-10 There was generally a greater burden of prognostic factors as the look back period increased. Across all of Sweden, the overall prevalence of individuals with at least one prognostic factor ranged from 19.4% with a one year look back to 27.6% with a ten year look back, and the overall prevalence of individuals with at least three prognostic factors ranged from 0.5% with a one year look back to 2.9% with a ten year look back (Appendix 2).

## DISCUSSION

Using data from the whole Swedish population, we show that over 2 million individuals (22.1%) have at least one of six prognostic factors for severe COVID-19 disease if infected, as defined by the World Health Organization and the European Centre for Disease Prevention and Control (cardiovascular disease, cancer, COPD, severe asthma, diabetes, or age 70 years and older). More than 150,000 individuals (1.6%) have at least three of these prognostic factors, which identifies the most vulnerable population. We also show that the distribution of the prognostic factors is heterogeneous across Sweden, with the Kalmar county containing the highest proportion of its inhabitants with at least one factor (25.5%). However, due to its high population density in comparison with other counties, Stockholm county has the highest number of individuals with at least one prognostic factors (416,988 individuals). We also present age and county specific prevalence of each prognostic factor to facilitate capacity planning and to provide underlying data for assumptions made in mathematical modelling of the current pandemic.

### Comparison to other studies

The number of people living with cardiovascular disease in 2015 in Sweden was 492,943 according to the Global Burden of Diseases studies, which is higher than the 389,774 we reported in the one year look back estimate in Appendix 2.[18] However, previous studies suggest the specificity and sensitivity of diagnoses of specific cardiovascular diagnoses such as acute myocardial infarction, heart failure and atrial fibrillation from the National Patient Registers are high.[19] The National Cancer Register reported that there were 214,000 individual tumors reported in 3 years prior to 31^st^ December 2016, while we report 129, 155 individuals in our study population with at least one tumor in 3 years prior to 1^st^ January 2016.[20] The prevalence of COPD is estimated at around 4-10% in Sweden, which is higher than the 0.8% we calculated. However, only 30% of COPD cases are diagnosed by healthcare professionals, which are often the most severe cases. The low observed prevalence of COPD in our study may therefore be due to us only capturing severe disease that is recorded in the Patient Register, and we were unable to capture moderate and mild COPD diagnosed and treated exclusively in primary care.[21-23] We have also underestimated the prevalence of asthma in Sweden, which is known to be between 8-10%, as we were only able to identify severe cases that required hospital admission.[19, 24, 25] However, more severe asthma is likely to exacerbate more severe COVID-19 disease, meaning we have identified those at the greatest risk. Finally, a study calculated a diabetes prevalence of 4.6% in Stockholm using survey data from the Stockholm Public Health Cohort, which is slightly higher than our estimate of 4.0%.[26] However, a report from the National Diabetes Register suggests that 22.6% of diabetes cases do not require pharmaceutical therapies, and only can be identified from primary care or quality registers, for which we did not have access.[27, 28]

### Strengths and limitations

Sweden is one of the few countries in the world where the study population for an analysis is the whole country. It is therefore possible to accurately calculate prevalence of underlying medical conditions for the whole population, without sampling.

We were only able to identify the burden of prognostic factors on 1^st^ January 2016. However, it is unlikely that the structure of the Swedish population has changed enough in four years to considerably change the prevalence estimates we calculated. The population of Sweden has increased by 476,572 inhabitants between 1^st^ January 2016 and 1^st^ January 2020.[29]

We could not identify all underlying medical conditions that the World Health Organization and the European Centre for Disease Prevention and Control suggest are prognostic factors for severe COVID-19 disease. Given the data we had available, we were not able to identify individuals with hypertension or high blood pressure because these conditions are usually diagnosed in primary care, and we only had access to data from specialized outpatient care and hospitalizations.[30] The Prescribed Drug Register could identify individuals with hypertension or high blood pressure as it includes information on individuals that dispensed a medication regularly used to treat these conditions (diuretics, beta-blockers, ACE inhibitors etc.).[30] However, the data we had available from the Prescribed Drug Register did not include information on these medications. Given hypertension and high blood pressure are precursors of clinical cardiovascular disease, it is likely that those with the most severe disease are captured in our cardiovascular disease estimates. Furthermore, other health agencies around the world (Center for Disease Control, United States; National Health Service, United Kingdom) have suggested additional prognostic factors for severe COVID-19 disease such as chronic kidney disease, liver disease, immunosuppression, and severe obesity. We decided to ground our choice of prognostic factors on recommendations from the World Health Organization and the European Centre for Disease Prevention and Control to give an overview of factors deemed important by multinational organizations, and as these are likely the guidelines that individuals in Sweden and Europe are currently following.

There is little available evidence on the prognostic factors that contribute the most to severe COVID-19 disease in different populations across the world. Our raw measure of cumulative number of prognostic factors for severe COVID-19 disease may therefore not represent those at the highest risk if one factor contributes more to severe disease in comparison with the others. Data from the World Health organization-China Joint Mission on Coronavirus Disease suggest that the case-fatality is highest in those with cardiovascular disease (13.2%) compared with cancer (7.6%), chronic respiratory disease (8.0%), diabetes (9.2%), and those with no comorbid conditions (1.4%).[31] If this is similar in all populations, then the individual prevalence of each of the prognostic factors at national and county level in Sweden that we have also presented may give better information of the populations at highest risk of severe COVID-19 disease.

All calculations from the main analyses rest on the assumption that any medical conditions were diagnosed within three years prior to 1^st^ January 2016. We have also presented the same analyses when the look back period is changed to one, five and ten years. The primary look back period was defined as three years due to being a reasonable time frame to capture individuals with active disease. We believe that this is a fair assumption and gives an accurate overview of the burden of disease in the population for all diseases apart from cancer. It has been suggested that only those with active cancer are truly at a high risk of severe COVID-19, and a definition of active cancer can take many forms.[32] Three years can be considered a long time after cancer diagnosis, and if the individual has survived, it is likely they will be considered to no longer have active cancer at three years after diagnosis. Therefore, for cancer, the analysis with a one year look back period may be a better estimation of individuals with active disease.

This study gives an accurate overview of the burden and prevalence of individuals in Sweden with the prognostic factors for severe COVID-19 disease. We have not made any attempt to model the transmission of the disease, but rather provide clear calculations of the number of vulnerable individuals based on current guidelines. The burden of severe COVID-19 disease will not reach the numbers we report in this study if public health interventions and mitigation strategies are successful. However, these numbers will allow authorities to optimally plan healthcare resources, by comparing the number of individuals at risk of severe disease with the critical care capacity. These results can also be applied to underlying assumptions of disease burden in modelling efforts to support COVID-19 planning. Overall, this information is crucial when deciding appropriate strategies to mitigate the pandemic and reduce both the direct mortality burden from the disease itself, and the indirect mortality burden from potentially overwhelmed health systems.

## Data Availability

All data are freely available within the manuscript and appendices. No additional data available.

## FOOTNOTES

The Corresponding Author has the right to grant on behalf of all authors and does grant on behalf of all authors, a worldwide licence to the Publishers and its licensees in perpetuity, in all forms, formats and media (whether known now or created in the future), to i) publish, reproduce, distribute, display and store the Contribution, ii) translate the Contribution into other languages, create adaptations, reprints, include within collections and create summaries, extracts and/or, abstracts of the Contribution, iii) create any other derivative work(s) based on the Contribution, iv) to exploit all subsidiary rights in the Contribution, v) the inclusion of electronic links from the Contribution to third party material where-ever it may be located; and, vi) licence any third party to do any or all of the above.

### Contributors

AM and KG generated the project idea. All authors contributed to the study design. AM and MT carried out all analyses. AM and KG drafted the manuscript. All authors contributed to further drafts and approved the final manuscript. AM is guarantor.

### Funding

There was no specific funding for this project.

### Conflicts of interest

All authors have nothing to disclose.

### Ethical approval

Approved by the Regional Ethical Review Board in Stockholm (2011/634-31/4, 2016/27-32, 2018/1257-32).

### Data sharing

All data are freely available within the manuscript and appendices. No additional data available.

### Transparency

The lead author affirms that the manuscript is an honest, accurate, and transparent account of the study being reported; that no important aspects of the study have been omitted; and that any discrepancies from the study as planned (and, if relevant, registered) have been explained.

## WHAT THIS PAPER ADDS

### What is already known on this topic

- The World Health Organization and the European Centre for Disease Prevention and Control suggest that individuals over the age of 70 years or with underlying cardiovascular disease, cancer, chronic obstructive pulmonary disease, asthma, or diabetes are at increased risk of severe COVID-19 disease.
- Governments around the world now recommend that individuals with at least one of these prognostic factors for severe COVID-19 self-isolate for prolonged periods of time to not only reduce the risk contracting severe disease, but also prevent any sudden increase in demand for critical care in hospitals, which could overwhelm health systems.

### What this study adds

- This study reports that over 22% of the Swedish population have at least one of the prognostic factors, and 1.6% have at least three factors.
- The prevalence of prognostic factors is heterogenous across Sweden, with the Kalmar county containing the highest proportion of its inhabitants with at least one factor (25.5%). However, due to its high population density, Stockholm county has the highest number of individuals with at least one prognostic factor (416,988 individuals).
- These figures can be used to aid healthcare capacity planning to overcome any shortage in critical care beds and contribute towards underlying assumptions made in modelling efforts to support COVID-19 planning.

## Notes

### Competing Interest Statement

The authors have declared no competing interest.

### Clinical Trial

This is a register based observational study, using only information from administrative health care registers, it requires no inform consents and registration

